# Environmental Risk Factors Influence the Natural History of Familial Dilated Cardiomyopathy

**DOI:** 10.1101/2024.06.25.24309501

**Authors:** Stacey A. Peters, Leah Wright, Jess Yao, Lauren McCall, Tina Thompson, Bryony Thompson, Renee Johnson, Quan Huynh, Celine F Santiago, Alison Trainer, Mark Perrin, Paul James, Dominica Zentner, Jon Kalman, Thomas H. Marwick, Diane Fatkin

## Abstract

**Background:** Familial dilated cardiomyopathy (DCM) is characterized by marked variability in phenotypic penetrance. The extent to which this is determined by patient-specific environmental factors is unknown.

**Methods:** A retrospective longitudinal cohort study was performed in families with DCM-causing genetic variants. Environmental factors were classified into two subsets based on evidence for a causal link to depressed myocardial contractility, termed (1) DCM-promoting factors and (2) heart failure (HF) comorbidities. These factors were correlated with DCM diagnosis, disease trajectory, and adverse events.

**Results:** 105 probands and family members were recruited: 51 genotype-positive, phenotype-positive (G+P+), 24 genotype-positive, phenotype-negative (G+P-), and 30 genotype-negative, phenotype-negative (G-P-). Baseline characteristics were similar between the 3 genotype groups. DCM-promoting environmental factors (eg. alcohol excess) were enriched in G+P+ individuals compared to G+P-(*P*<0.001) and G-P-(*P*=0.003) and were significantly associated with age at DCM onset (HR 2.01, *P*=0.014). HF comorbidities (eg. Diabetes) had a similar prevalence in G+P+ and G-P-but were significantly reduced in the G+P-group. Fluctuations in left ventricular ejection fraction during follow-up were linked to changes in environmental factors in 35/45 (78%) of instances: 32 (91%) of these were DCM-promoting factors. HF comorbidities, but not DCM-promoting factors, were associated with adverse events in G+ individuals (OR 4.9, *P*=0.004).

**Conclusion:** We identified distinct subsets of environmental factors that affect DCM penetrance and adverse outcomes respectively. Our data highlight DCM-promoting environmental factors as key determinants of penetrance and disease trajectory. Collectively, these findings provide a new framework for risk factor assessment in familial DCM and have important implications for clinical management.

## Introduction

Dilated cardiomyopathy (DCM) is an important heart muscle disease associated with significant morbidity and mortality from heart failure (HF), arrhythmias, and sudden cardiac death.^1^ DCM can arise as a primary myocardial disorder or as a consequence of diverse endogenous and exogenous factors that impair myocardial function. Inherited gene sequence variants are frequently responsible for primary forms of DCM. Approximately 30% of DCM is familial, and genetic testing identifies a single disease-causing (i.e., pathogenic [P] or likely-pathogenic [LP]) variant in 30-40% of familial cases.^1,2^ Studies in cohorts of probands with P/LP variants have sought to identify gene-specific phenotypic features; however, there is often considerable variability in the penetrance and severity of DCM associated with variants in a single gene or in family members with the same variant, and this makes clinical management challenging.

Phenotypic variability in genetic forms of DCM may be determined, at least in part, by the patient-specific context.^3,4^ This encompasses background genetic variation and a range of physiological conditions, comorbidities, lifestyle choices, and exogenous factors that influence the myocardial environment (see Box). There are emerging data to suggest that patients with genetically-mediated DCM have a greater arrhythmia burden and worse outcomes if there are concurrent environmental risk factors.^5,6^ A number of key questions remain, however, including what the most important environmental risk factors are, the impact of acute vs chronic exposure on disease trajectory in genotype-positive individuals with and without established DCM, and reversibility of effects.

### Box

| KEY DEFINITIONS |
| --- |
| **Myocardial environmental factors:** |
| Patient-specific combinations of physiological conditions, comorbidities, lifestyle patterns and exogenous agents that influence the myocardial environment and ventricular contractile function. |
| **DCM-promoting risk factors:** |
| Endogenous or exogenous factors that can depress ventricular contractile function and promote DCM via direct toxic, mechanical, inflammatory, or metabolic effects. |
| **Heart failure comorbidities:** |
| Endogenous or exogenous factors associated with heart failure but not specifically with DCM. |
| **Phenotype plasticity:** |
| Window in which ventricular function retains the capacity to be reversibly altered by environmental exposures. |

The purpose of this study was to report a retrospective longitudinal cohort study to investigate associations between environmental risk factors and phenotypic manifestations of familial DCM.

## Methods

### Study Cohort

The study cohort comprised adults referred to the Royal Melbourne Hospital Cardiac Genetics Clinic between 2009 and July 2022 for assessment of suspected familial DCM. Genetic testing was performed by NATA accredited clinical laboratories and probands found to carry P/LP variants were selected for potential inclusion. All variants were re-assessed by the research team using modified ACMG/AMP criteria.^7^ Probands with variants that were re-classified as variants of uncertain significance or that were in genes deemed to have low evidence for DCM pathogenicity based on updated ClinGen gene curation were excluded.^8^ Eligible probands and their relatives were contacted by phone and invited to participate in the study. Participants were retrospectively followed from their first clinical echocardiogram to the study end in July 2022. All living participants provided informed written consent and protocols were approved by the Melbourne Health ethics review board (HREC/18/MH/117). Data on deceased individuals were included if another family member consented on their behalf.

### Clinical Information

At the time of enrolment, participants underwent a comprehensive phone or in-person interview and health record interrogation. Data collected included: demographics, medical history, physical examination, and a three-generation family pedigree. Information on deceased individuals was gathered from relatives and medical records.

### Environmental Factors

A list of putative environmental risk factors associated with DCM or HF was sourced from the literature.^9–13^ Each factor was then reviewed in detail and a semiquantitative grading method was applied to indicate the level of supportive evidence for disease association see Table 2 and footnote). Parameters considered included numbers of reported cases, clinical trial evaluation, and experimental data. A pre-specified coding system was derived to define the level of exposure in individual study subjects (Supplemental Table 1).

**Table 1.**
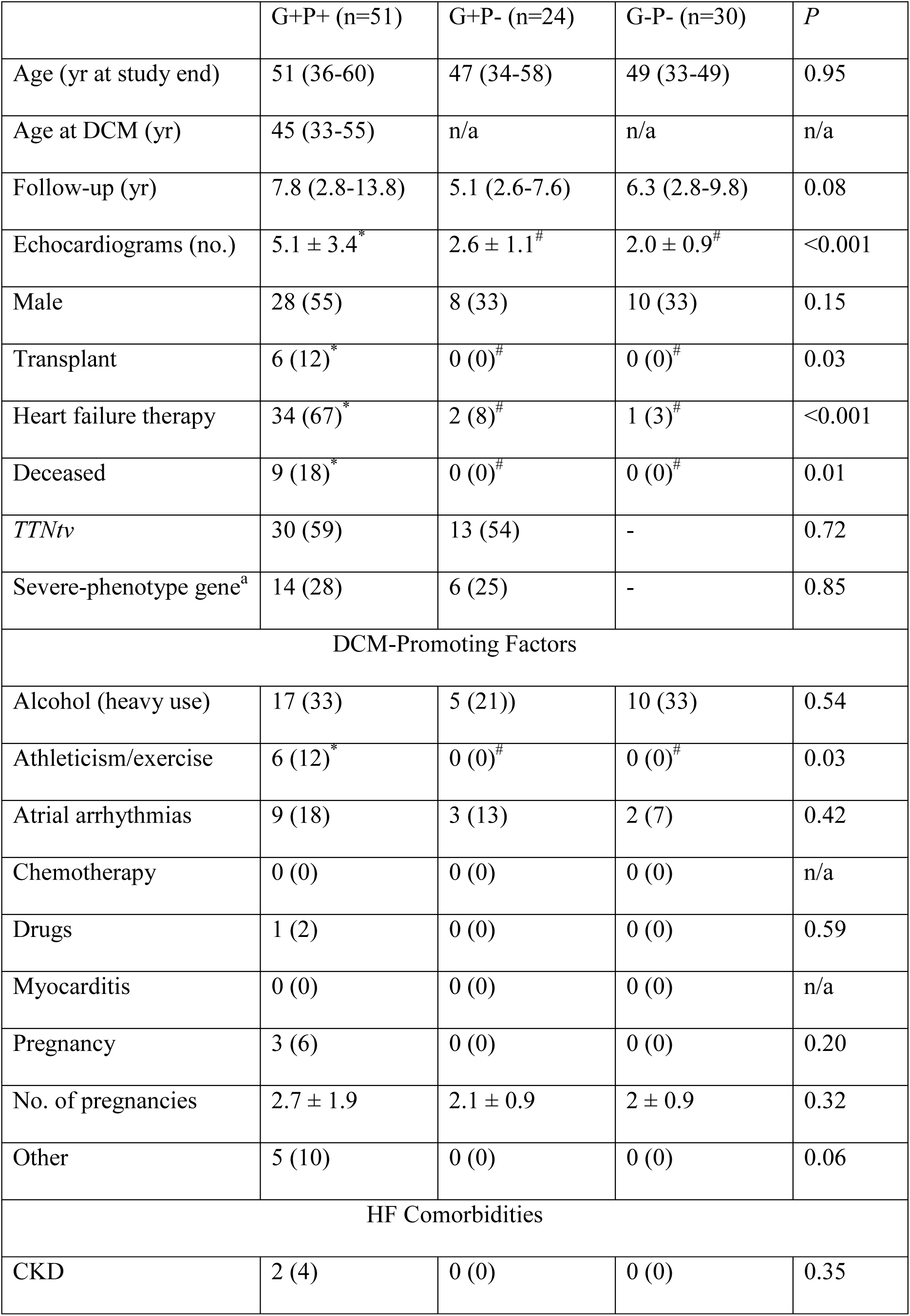
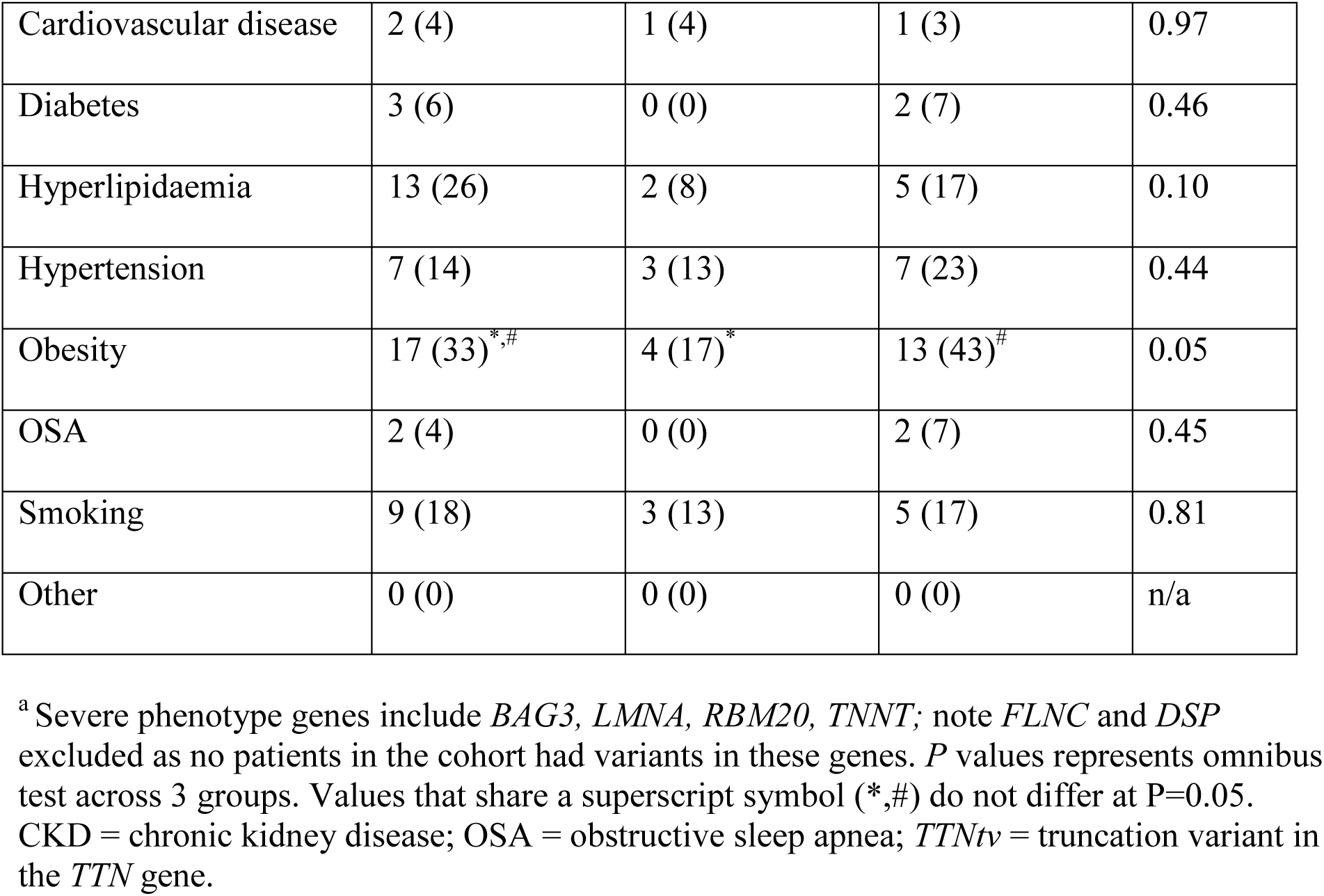
Baseline Characteristics of the Cohort.

|  | G+P+ (n=51) | G+P- (n=24) | G-P- (n=30) | *P* |
| --- | --- | --- | --- | --- |
| Age (yr at study end) | 51 (36-60) | 47 (34-58) | 49 (33-49) | 0.95 |
| Age at DCM (yr) | 45 (33-55) | n/a | n/a | n/a |
| Follow-up (yr) | 7.8 (2.8-13.8) | 5.1 (2.6-7.6) | 6.3 (2.8-9.8) | 0.08 |
| Echocardiograms (no.) | 5.1 ± 3.4<sup>*</sup> | 2.6 ± 1.1<sup>#</sup> | 2.0 ± 0.9<sup>#</sup> | <0.001 |
| Male | 28 (55) | 8 (33) | 10 (33) | 0.15 |
| Transplant | 6 (12)<sup>*</sup> | 0 (0)<sup>#</sup> | 0 (0)<sup>#</sup> | 0.03 |
| Heart failure therapy | 34 (67)<sup>*</sup> | 2 (8)<sup>#</sup> | 1 (3)<sup>#</sup> | <0.001 |
| Deceased | 9 (18)<sup>*</sup> | 0 (0)<sup>#</sup> | 0 (0)<sup>#</sup> | 0.01 |
| *TTNtv* | 30 (59) | 13 (54) | - | 0.72 |
| Severe-phenotype gene<sup>a</sup> | 14 (28) | 6 (25) | - | 0.85 |
| DCM-Promoting Factors |  |  |  |  |
| Alcohol (heavy use) | 17 (33) | 5 (21)) | 10 (33) | 0.54 |
| Athleticism/exercise | 6 (12)<sup>*</sup> | 0 (0)<sup>#</sup> | 0 (0)<sup>#</sup> | 0.03 |
| Atrial arrhythmias | 9 (18) | 3 (13) | 2 (7) | 0.42 |
| Chemotherapy | 0 (0) | 0 (0) | 0 (0) | n/a |
| Drugs | 1 (2) | 0 (0) | 0 (0) | 0.59 |
| Myocarditis | 0 (0) | 0 (0) | 0 (0) | n/a |
| Pregnancy | 3 (6) | 0 (0) | 0 (0) | 0.20 |
| No. of pregnancies | 2.7 ± 1.9 | 2.1 ± 0.9 | 2 ± 0.9 | 0.32 |
| Other | 5 (10) | 0 (0) | 0 (0) | 0.06 |
| HF Comorbidities |  |  |  |  |
| CKD | 2 (4) | 0 (0) | 0 (0) | 0.35 |
| Cardiovascular disease | 2 (4) | 1 (4) | 1 (3) | 0.97 |
| Diabetes | 3 (6) | 0 (0) | 2 (7) | 0.46 |
| Hyperlipidaemia | 13 (26) | 2 (8) | 5 (17) | 0.10 |
| Hypertension | 7 (14) | 3 (13) | 7 (23) | 0.44 |
| Obesity | 17 (33)<sup>*,#</sup> | 4 (17)<sup>*</sup> | 13 (43)<sup>#</sup> | 0.05 |
| OSA | 2 (4) | 0 (0) | 2 (7) | 0.45 |
| Smoking | 9 (18) | 3 (13) | 5 (17) | 0.81 |
| Other | 0 (0) | 0 (0) | 0 (0) | n/a |

<sup>a</sup> Severe phenotype genes include *BAG3*, *LMNA*, *RBM20*, *TNNT*; note *FLNC* and *DSP* excluded as no patients in the cohort had variants in these genes. *P* values represents omnibus test across 3 groups. Values that share a superscript symbol (\*,#) do not differ at $P=0.05$ . CKD = chronic kidney disease; OSA = obstructive sleep apnea; *TTN*<sub>tv</sub> = truncation variant in the *TTN* gene.

**Table 2.**
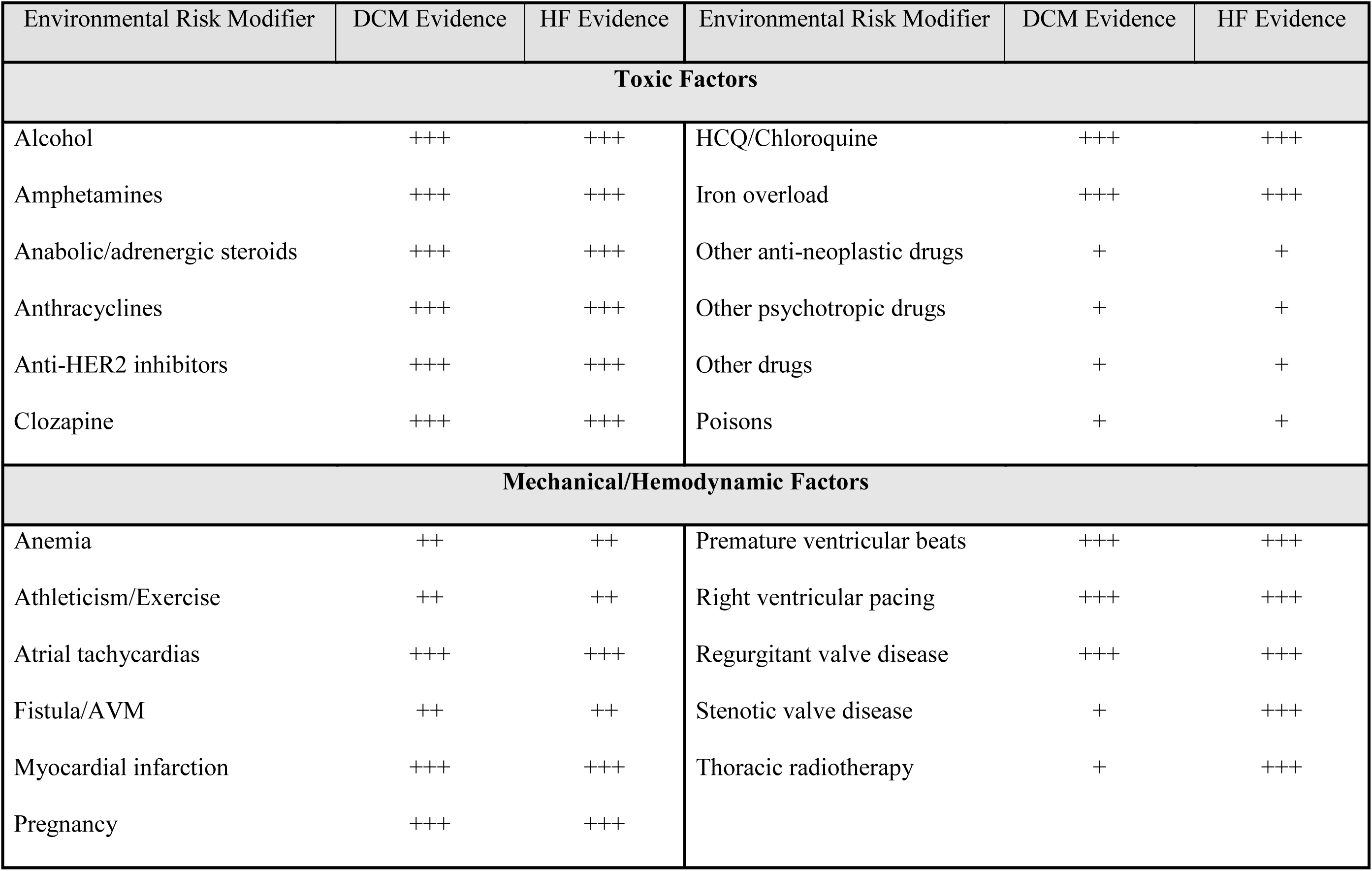
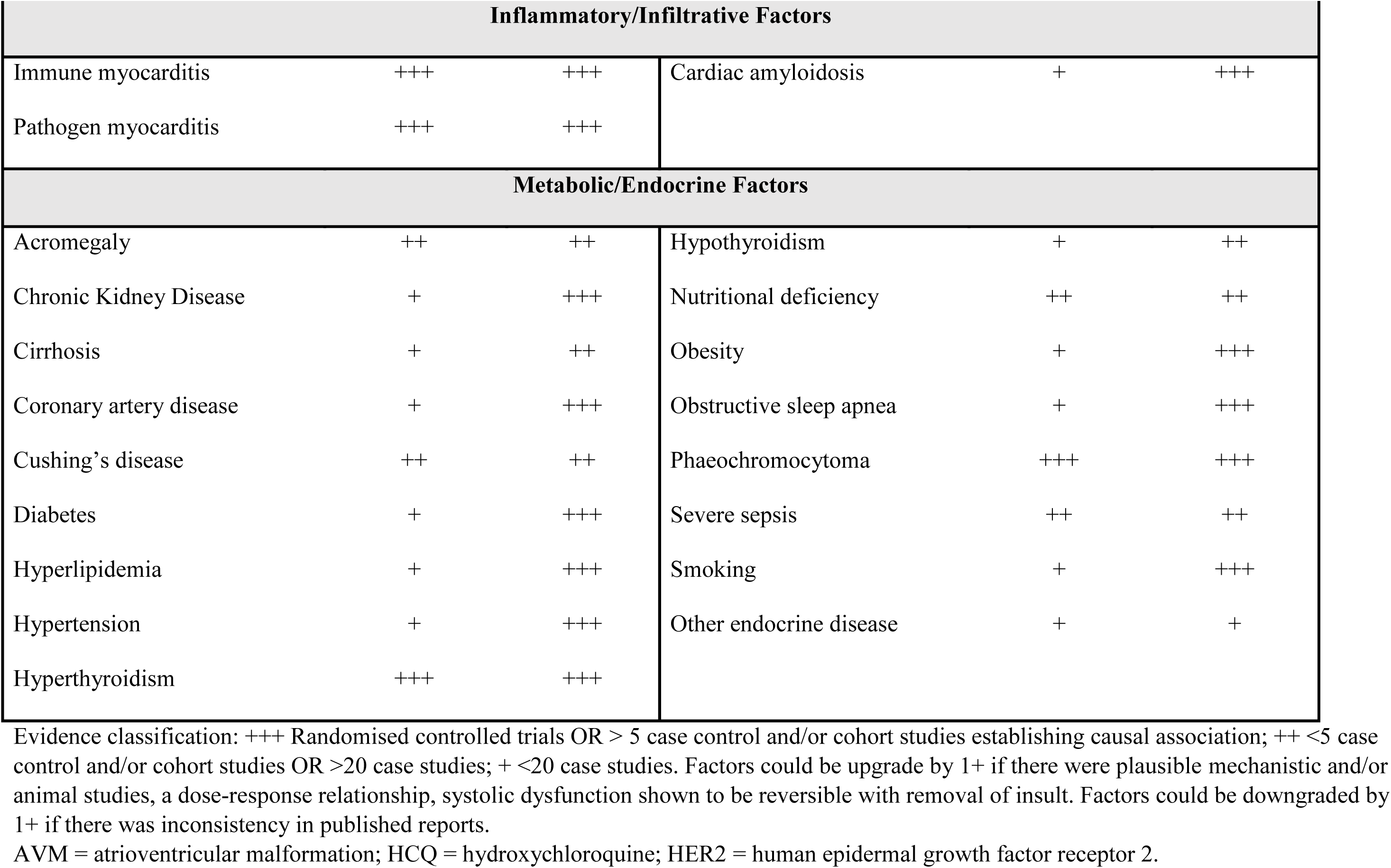
Environmental Risk Factors Stratified by Mechanistic Category.

### Transthoracic Echocardiography

Participants underwent serial transthoracic echocardiography as part of their routine medical management. These studies were performed at various sites and reports were gathered for the baseline echocardiogram and for subsequent echocardiograms undertaken during the follow-up period. For quality control, a subset of available echocardiograms from the clinical sites was formally re-analyzed at a central laboratory (Baker Heart and Diabetes Institute). Images were downloaded to DICOM format and evaluated by a blinded experienced sonographer using third party software (Image Arena, TomTec Imaging Systems, Munich, Germany).

Conventional clinical echocardiographic parameters were measured according to recommendations of the American Society of Echocardiography.^14^ LV volumes and LV ejection fraction (LVEF) were calculated by the biplane method of disks. The intraclass correlation coefficient between measures of LV function obtained at clinical sites and the central laboratory was determined. Disease trajectory was considered changed if there was a difference (±) in absolute LVEF of >10% within 2 years (if at least one measurement included LVEF ≤50%). For mean LVEF curves, sequential echocardiograms were divided into time periods to create uniformity in the heterogenous duration of follow-up. Sequential time periods were: baseline, 1-6 months, 7-12 months, 1-3 years, 3-5 years, 5-10 years and > 10 years.

### Adverse Outcomes

Adverse cardiovascular events comprised a broad composite of: cardiovascular death, heart transplant, HF hospitalization or ventricular arrhythmias (ventricular fibrillation arrest, sustained ventricular tachycardia).

### Statistical Analysis

Categorical variables are reported as frequencies and percentages, and continuous variables as means ± standard deviation or median (interquartile range) as appropriate. Where there were missing data points for particular risk factors, they were coded as absent. For all comparisons, differences between categorical variables were assessed using logistic regression and continuous variables using general linear models. Adjustment for multiple comparisons between groups was conducted using a post-hoc Bonferroni correction where applicable. When categories included a group with zero counts, one-way ANOVA or independent samples T tests were used. For all analyses, cluster robust standard errors were used to account for within-family dependencies. To assess age at onset of DCM, a time-to-event analysis was performed with the use of a Cox proportional hazards model with time to endpoint being age at DCM diagnosis or censoring at study end (July 2022). Factors and covariates included in regression models included: age (at DCM diagnosis or at time of study if no DCM), sex, disease gene associated with severe phenotypes (*BAG3, DSP, FLNC, LMNA, RBM20, TNNT2*), and number of environmental factors. Test of proportionality were met for each covariate. An alpha level of 0.05 was considered significant, though due to the multiple comparisons within the study all results are considered hypothesis generating. All analyses were carried out using SPSS version 28 (IBM Corp., Armonk, NY, USA).

## Results

### Genotyped Families

128 probands with familial DCM were referred for genetic testing: 39 (30%) probands were found to have a P/LP variant (Figure 1). Of these, 8 probands were unable to be recontacted. The final study group comprised 105 genotyped individuals (median age 48 years [interquartile range 35 - 60], 56% female), from 31 families (median 2 [range 1-16] individuals per family (Figure 1). The majority of participants (99%) were of self-reported Western European ancestry. 51 genotype-positive (G+) individuals with DCM were classified as phenotype-positive (G+P+), 24 G+ individuals were phenotype-negative (G+P-) and 30 screened unaffected individuals were genotype-negative (G-P-) (Figure 1). There were no significant differences in age or gender between the three groups (Table 1). Seventeen families (55%) had truncating variants in *TTN (TTN*tv), while the remainder had variants in *LMNA* (n=3), *PLN* (n=2)*, TNNT2* (n=2), *RBM20* (n=2), *DES* (n=2), *BAG3* (n=1), *MYH7* (n=1) and *SCN5A* (n=1). Three variants, one each in *PLN, TNNT2* and *TTN*, were identified in two unrelated probands. No individuals had more than one clinically important variant. Variant details are shown in Supplemental Table 2.

**Figure 1.**
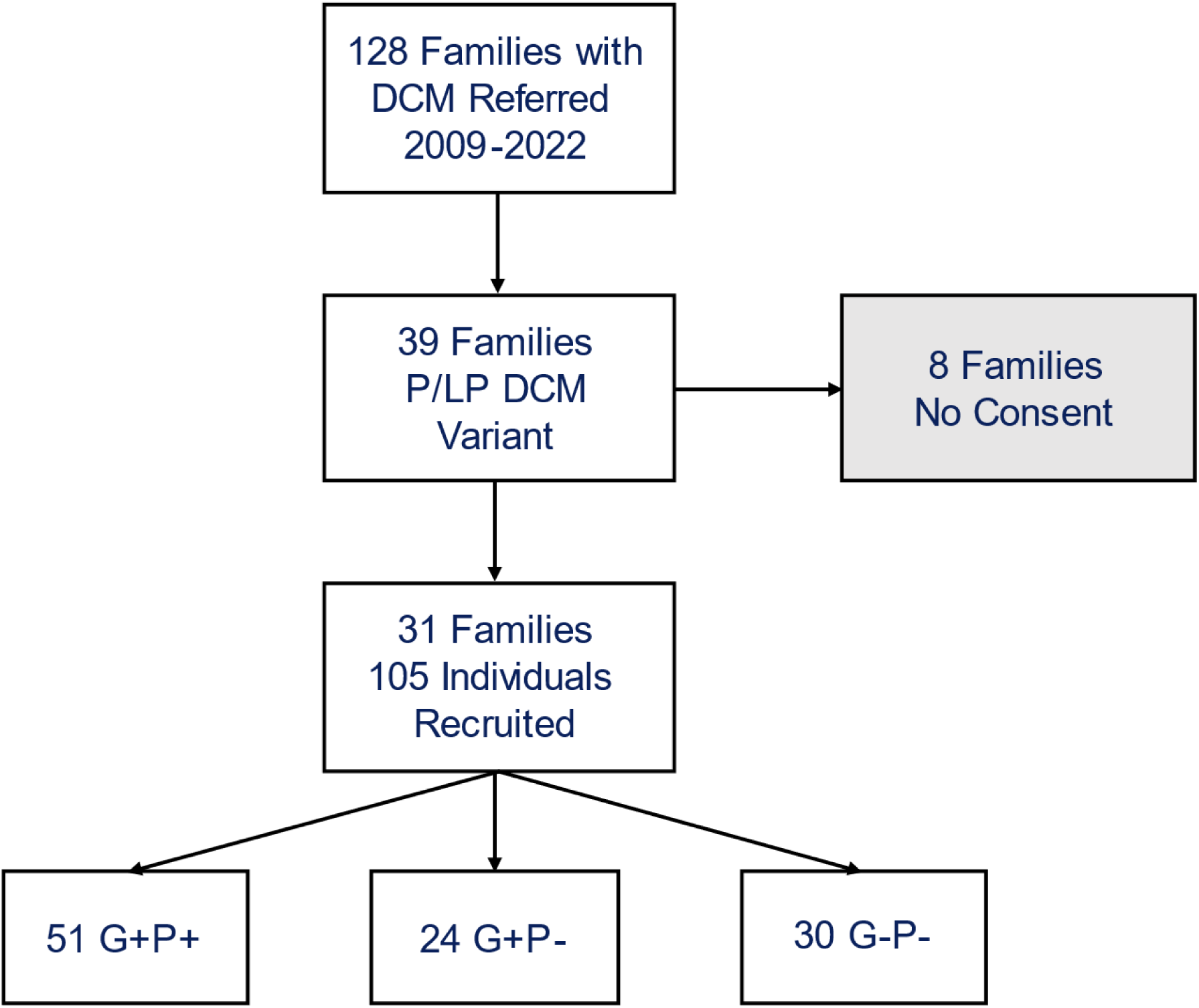
Recruitment Flowchart. Recruitment flowchart for the derivation of the study cohort. DCM, dilated cardiomyopathy; G+P+, genotype-positive phenotype-positive; G+P-, genotype-positive phenotype-negative; G-P-, genotype-negative (phenotype-negative); P/LP = pathogenic or likely-pathogenic DCM genetic variant.

### Identification of Distinct Environmental Risk Factor Subtypes

Forty-three environmental factors associated with heart failure were identified from the literature search (Table 2). Twenty-five factors had moderate or strong evidence for an association with DCM and HF, these were termed *DCM-promoting factors* (see Box). A second group of 13 factors had moderate or strong evidence for HF but low evidence of DCM, and these were termed *HF comorbidities* (see Box). A further five factors had low evidence for both DCM and HF and were excluded. DCM-promoting factors included conditions that can depress myocardial function via direct toxic effects (e.g. alcohol excess, anthracycline chemotherapy, drug misuse), mechanical stress (e.g. persistent tachycardia, pregnancy), metabolic or endocrinological disturbance (e.g. hyperthyroidism), or inflammation (e.g. viral infection). HF comorbidities included cardiovascular and metabolic disorders such as hypertension, diabetes, and obesity. For statistical analyses, factors within each of the DCM-promoting and HF comorbidity groups were collapsed into 8 or 9 subgroups, respectively (Table 3).

**Table 3.** Collapsed List of Environmental Factors Included in the Final Analysis.

| **DCM-Promoting Factors** | **HF Comorbidities** |
| --- | --- |
| Alcohol | Chronic kidney disease |
| Athleticism/exercise | Coronary artery disease |
| Atrial tachyarrhythmias | Diabetes |
| Chemotherapy | Hyperlipidaemia |
| Drug misuse | Hypertension |
| Myocarditis | Obesity |
| Pregnancy | Obstructive sleep apnea |
| Other | Smoking |
|  | Other |

### Role of DCM-Promoting Environmental Risk Factors

The relationship between DCM-promoting environmental factors (listed in Table 3) and DCM onset in G+ individuals was determined (Figure 2). After adjustment for covariates, the mean number of DCM-promoting factors was significantly higher in G+P+ individuals (0.84±0.73) when compared to both G+P-(0.29±0.46, *P*<0.001) and G-P-(0.43±0.57, *P*=0.003; Figure 2A). The most commonly encountered factors were heavy alcohol use and atrial arrhythmias, however other factors noted at DCM diagnosis included pregnancy, high levels of exercise (>12hrs/week), amphetamine use, and new right ventricular pacing (>70%). In a time-to-event model, the presence of a DCM-promoting factor was significantly associated with age at onset of DCM (HR 2.01, 95% CI 1.17-3.47, *P*=0.014; Figure 2B).

**Figure 2.**
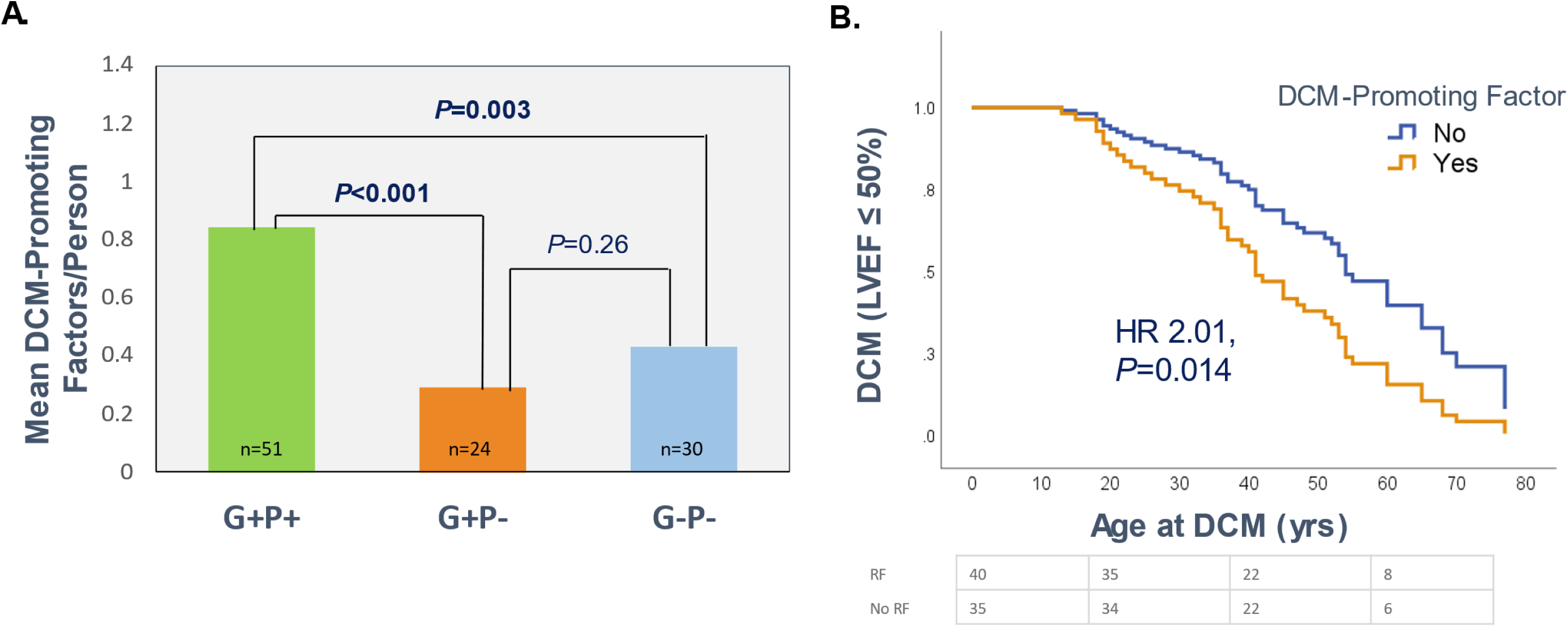
Association between DCM-Promoting Environmental Risk Factors and DCM Onset in Genotyped Individuals. **(A)** Bar graph comparing the mean number of DCM-promoting factors per person in each of the three genotype groups. Data were analyzed using a general linear model with adjustment for covariates (age, sex, and severe-phenotype gene). **(B)** Cox proportional hazards curve demonstrating survival free from DCM in G+ individuals stratified into those with and without DCM-promoting factors.

### Role of Heart Failure Comorbidities

HF comorbidities (listed in Table 3) were similarly prevalent in G+P+ (mean number of factors, 0.84 ± 0.73) and G-P-individuals (1.2 ± 1.3, *P*=0.41; Figure 3A). These findings indicate that the presence of HF comorbidities alone was insufficient to cause DCM. In contrast, G+P-individuals had significantly fewer HF comorbidities (0.29 ± 0.46) when compared to both G+P+ (*P*=0.003) and G-P-(*P*=0.01). It is unclear whether the lower number of HF comorbidities *per se*, or other health-promoting activities that mitigate against the development of these risk factors, might account for the absence of DCM in this subgroup. The presence of any HF comorbidity was not significantly associated with age at onset of DCM in a Cox regression model (HR 0.76, 95% CI 0.44-1.29, *P*=0.29; Figure 3B).

**Figure 3.**
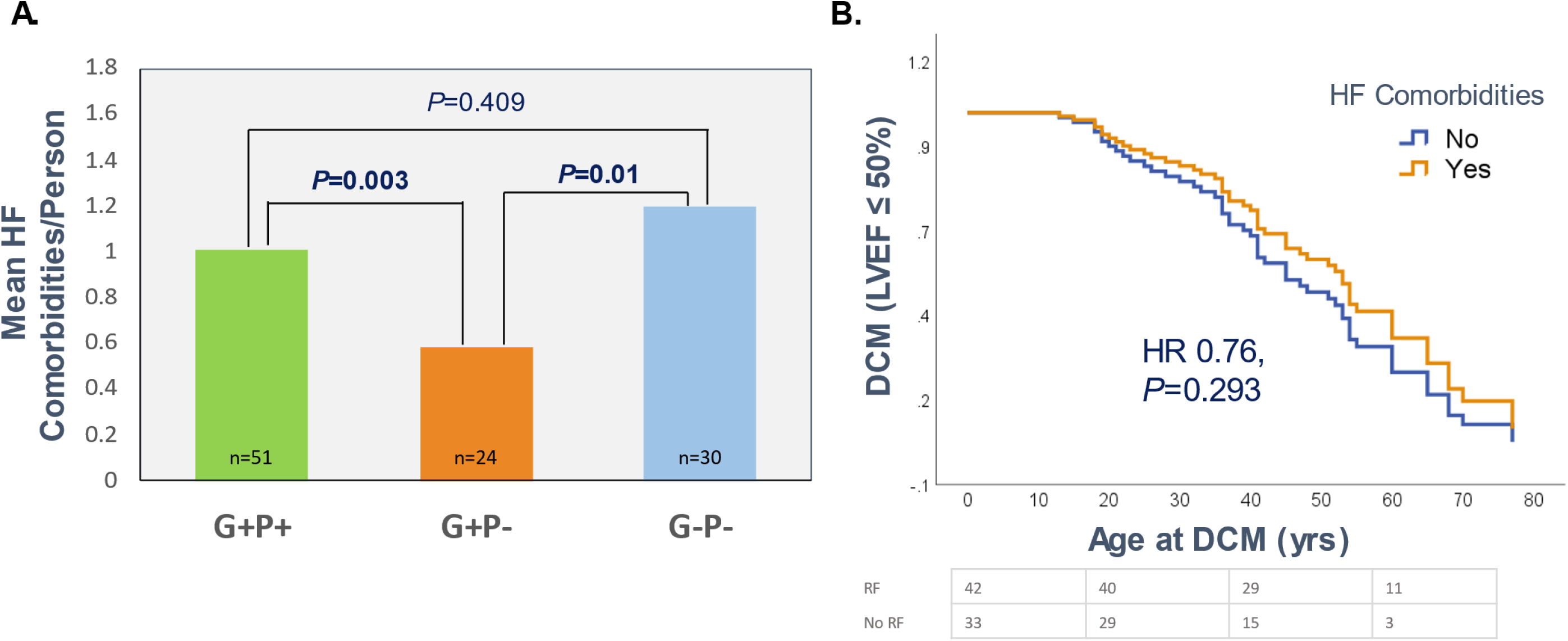
Association between HF Comorbidities and DCM Onset in Genotyped Individuals. **(A)** Bar graph comparing the mean number of HF comorbidities per person in three genotype groups. Data were analyzed using a general linear model with adjustment for covariates (age, sex, and severe-phenotype gene). **B)** Cox proportional hazards curve demonstrating survival free from DCM in G+ individuals with and without HF comorbidities.

### Natural History of LV Function in Genetically Susceptible Individuals

Disease trajectory in G+ individuals was determined by evaluation of LVEF reported on serial echocardiograms. These were undertaken at various clinical sites at the discretion of each individual’s usual practitioner. However, images from a subset of 89 individuals were available for additional standardized review at the central laboratory. There was good agreement between reported LVEF and biplane EF measured in the central laboratory with an intraclass correlation coefficient 0.82 (95% CI 0.74-0.89, *P*<0.001), suggesting that reported LVEF was a reasonably reliable indicator of LV function in these patients. On their baseline echocardiography report, 23 G+ individuals had LVEF <40%, 12 had LVEF between 40-50% and 40 had LVEF >50% (Figure 4A). Of these 75 G+ individuals, 65 had echocardiographic follow-up over a median 6.9 (2.9-10.9) years; serial data were not available for 10 individuals due to heart transplantation or death (n=7), or no echocardiogram performed (n=3). All patients with baseline LVEF <40% were started on HF therapy, as were 5/12 (42%) of those with baseline LVEF 40-50%. Despite therapy, 4 individuals in the baseline LVEF <40% group failed to improve, while 3 individuals who initially had LVEF >50% had deterioration to <40% (Figure 4A). In contrast, the majority of G+ individuals (89%) remained stable in the low-normal to mildly abnormal range. This included 19 individuals (29%) who had LV reverse remodelling (LVRR) (Figure 4 A & B).

**Figure 4.**
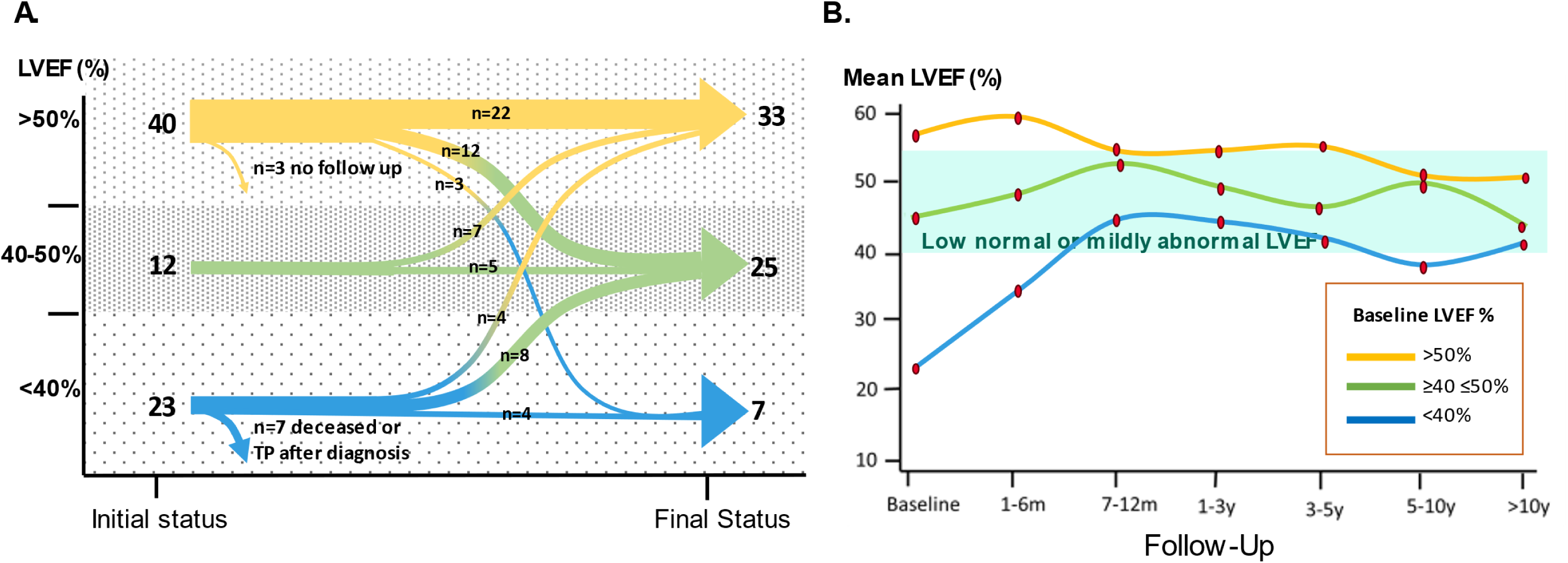
Disease Trajectory by Baseline LVEF. **(A)** Stratification of G+ individuals (n=75) into three groups based on baseline LVEF: <40%, 40-50% or >50%. Curves show overall movement between LVEF groups from baseline echo to the last available echo. **(B)** Mean LVEF trajectory curves in the three baseline LVEF groups. Note that data from the worst affected patients who died or were transplanted without follow up are not represented.

### Impact of Environmental Factors on LVEF Fluctuation

There were 45 instances of LVEF fluctuation (defined by absolute change in LVEF >10% within 2 years, with one measure ≤50%) in the G+ cohort, including 12 instances of deterioration and 23 instances of improvement (Figure 5). Thirty-five (78%) LVEF fluctuation episodes could be linked to a demonstrable change in an environmental factor: the majority of these (32 [91%]), were DCM-promoting factors, with only 3 (9%) being marked changes in a HF comorbidity. The most common factors associated with LVEF fluctuation were new heavy alcohol use (n=4) or alcohol abstinence (n=11), and new atrial arrhythmias (n=4) or atrial arrhythmia management (n=7). Other factors associated with subacute LVEF change included new high burden right ventricular pacing or cardiac resynchronization therapy, pregnancy delivery, acute myocardial infarction, treatment of severe undiagnosed hypertension, subacute severe weight gain (>25% body weight) and subsequent weight loss, and new severe hyperthyroidism and its management. Susceptibility to LVEF fluctuation differed between *TTN*tv and non-*TTN*tv genotypes. Notably, all *TTNtv* -positive individuals with reversible LVEF changes were found to have an attributable environmental factor (*TTN*tv, 19/19 vs. non-*TTN*tv, 6/11, *P*=0.001).

**Figure 5.**
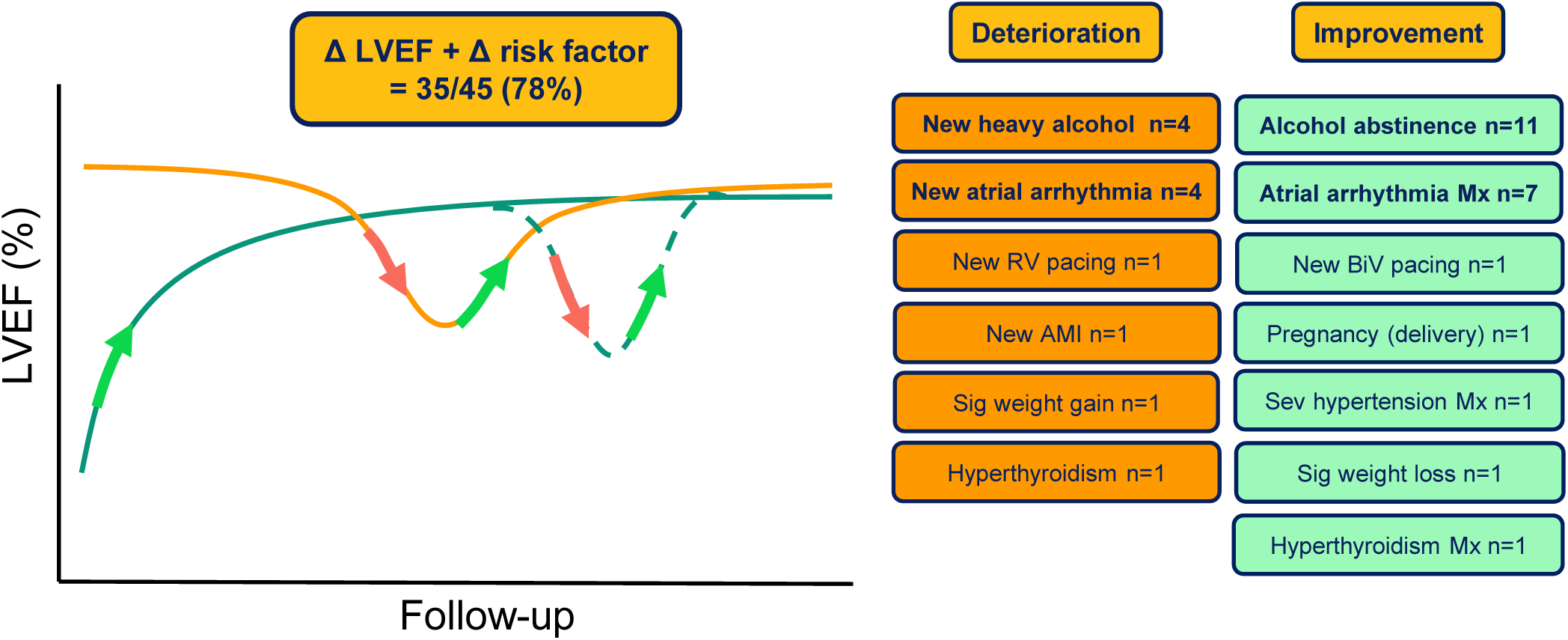
Fluctuations in LVEF and Environmental Factors. Schematic curve demonstrating changes in LVEF during follow-up in G+ individuals. Within the cohort, improvements and deteriorations >10% LVEF were seen, and 78% were linked to changes in environmental factors. Specific identified environmental factors that coincided with LVEF change episodes are listed; n= number of individuals impacted. AMI = acute myocardial infarction; BiV = biventricular; Mx = management; RV = right ventricular.

### Adverse Events

Adverse events occurred in 31 G+ individuals (41%), and no G-individuals. Stratified by baseline LVEF group, adverse events occurred in 22/23 (95%) with LVEF <40%, 3/12 (25%) with LVEF 40-50%, and 6/40 (15%) with LVEF >50%. There were no adverse events in individuals who remained G+P-at the end of the study. In univariate and multivariate analyses, only HF comorbidities and baseline LVEF, and not DCM-promoting factors, were independently associated with adverse events: HF comorbidities, OR 4.9, 95% CI 1.73-14.08, *P*=0.004; baseline EF, OR 1.26, 95% CI 1.15-1.39, *P*<0.001; DCM-promoting factors, OR 1.91, 95% CI 0.35-10.51, *P*=0.45 (Table 3). Ventricular arrhythmias or sudden cardiac death occurred in 18 individuals (24%) who had a mean LVEF of 37%. Concerningly, 8 events (44%) occurred in individuals with LVEF >40%: *TTN* (n=4), *LMNA* (n=1), *DES* (n=1), *SCN5A* (n=1), *TNNT2* (n=1). Of those with LVEF <40% at baseline, adverse events were common among those who did not respond to HF therapy, 10/11 died or underwent cardiac transplant during follow-up. However, among those who underwent LVRR, 9/12 had no adverse events, while three patients (*RBM20* [n=1], *TTN* [n=2]) had ventricular arrhythmias or sudden cardiac death.

## Discussion

Genotype-phenotype correlations in familial DCM have largely focused on genotype effects, with phenotypic differences often considered to be attributes of the underlying disease genes. It has been proposed that patient-specific factors that affect the myocardial milieu (i.e. myocardial environmental factors) might influence genetically-driven effects on ventricular function,^3^ but evidence for this has been lacking. Here we provide compelling new data that clearly define a role for environmental factors in the variable penetrance and natural history of familial DCM. We identified two main groups of environmental factors that affect DCM onset and outcomes, respectively. Moreover, we show that there is a vulnerable period in the DCM disease trajectory in which myocardial function can be reversibly altered by changes in environmental exposures. Collectively, our data have important clinical implications, highlighting the key role of risk factor surveillance in family management.

Based on literature evidence, we dichotomized environmental risk factors into two main groups, i.e., DCM-promoting factors and HF comorbidities. Differentiation between these two risk groups is highly relevant as there are distinctive etiologies, functional impacts, and consequences. The first group, DCM-promoting factors, can directly impair ventricular function. Accordingly, we found that DCM-promoting factors were relatively enriched in G+ individuals who had developed DCM. In our cohort, the most common DCM-promoting factors were alcohol excess and atrial tachyarrhythmias. Both of these conditions may contribute to a DCM substrate with chronic exposure or acutely trigger ventricular decompensation with new DCM diagnoses or HF episodes. Atrial arrhythmias were included as a DCM-promoting factor but are acknowledged to be part of the phenotypic spectrum of many DCM genotypes. Indeed, new evidence suggests that *TTN*tv produce an early tendency to atrial fibrillation.^6,15–17^ However, atrial arrhythmia was a common trigger for LV dysfunction in our cohort, and its management improved LV function, supporting its classification as a modifiable risk factor as well as a consequence of disease.

Heavy exercise (high level aerobic exercise >12 hours/wk) was associated with DCM in six individuals in this study. These relatively small numbers limit the conclusions that can be drawn, however reported observations of DCM in a subset of elite athletes raises the possibility of exercise-induced effects.^18^ Exercise recommendations in inherited cardiomyopathies have been debated.^19^ Until further research clarifies this point, it seems reasonable to recommend limitation of extreme exercise regimens on a case-by-case basis. Pregnancy was associated with DCM diagnosis in three women in this study, though pregnancy was common in the cohort generally with few retrospective complications noted (see Peters et al. for results of specific pregnancy analysis).^20^ Pregnancy is presumed to be an important trigger factor in familial DCM, but factors that promote this need further exploration.

There has been increasing interest in myocardial inflammation due to viral infection or other conditions as a precipitant of cardiomyopathic presentations.^21^ We did not identify any clear cases of myocarditis in this cohort. However, several studies have demonstrated the presence of subclinical inflammatory infiltrates and/or viral genomes in G+ patients on endomyocardial biopsy and these are hypothesized to promote disease.^5,6^ We did not perform endomyocardial biopsy or CMR routinely in this cohort, and hence some cases may have been missed. Further heart tissue studies are needed to clarify the role of immune-mediated and viral-mediated inflammation in DCM pathophysiology.

The impact of DCM-promoting factors may differ according to the underlying genotype and there may be additive or synergistic effects. As an example, *TTN*tv have been identified in ∼10% patients with alcohol-induced DCM.^22^ In recent studies from our group, we found that titin-deficient zebrafish hearts were more susceptible to myocardial depressant effects of alcohol than wildtype fish, indicating a gene-environment interaction.^23,24^ In this present study, DCM-promoting factors, particularly alcohol excess and atrial tachyarrhythmias, were uniformly present in *TTN*tv-positive individuals who experienced LVEF fluctuation. These findings might indicate that *TTN*tv hearts are particularly susceptible to toxic and mechanical stressors.

The second main group of environmental risk factors were conditions that have been associated more broadly with cardiovascular morbidity and mortality, including HF.^4^ Hypertension and obesity were the most common of these factors in our cohort. Unlike DCM-promoting factors, HF comorbidities were generally not associated with DCM penetrance but were associated with adverse events in G+ individuals. It is notable that HF comorbidities can on occasion, act like DCM-promoting factors. For example, treatment of severe uncontrolled hypertension or sudden marked weight gain/loss were associated with LVEF fluctuation in selected individuals, presumably due to marked changes in hemodynamic load and myocardial mechanical stress.

Among those with serial echocardiogram data, there was a trend to stabilization to low-normal or mildly abnormal LVEF in the majority of G+ individuals during a median 7 years of follow-up. Most patients were stabilized in the context of HF therapy. However, there were numerous instances of LVEF fluctuation that coincided with changes in an environmental factor. These data suggest that, rather than an inexorable decline in LV function over time, there may instead be a window of LV phenotypic plasticity, where cardiac function is vulnerable to the influence of environmental factors but can still be improved with HF therapy and/or modification of risk factors (See Box). How long is this window, what genetic and environmental factors predict its duration, and is there a point of no-return? Further longitudinal studies are required to address these questions.

A small group of G+ patients had preserved ventricular function throughout their follow-up. This was not explained by age or sex but was associated with a lower prevalence of DCM-promoting factors. It was interesting that while HF comorbidities were not clearly associated with penetrance in this study, they were also significantly lower in the G+P-subset. A focus on this group in future research may expand on factors that are more generally cardioprotective including exercise levels, dietary factors, medications or low polygenic risk scores for cardiomyopathy and other heart diseases.

Ongoing surveillance of G+ patients (regardless of baseline LVEF) is important and attention to reducing the burden of environmental risk modifiers is prudent. Surveillance of LVEF during pregnancy, chemotherapy, or other unavoidable potential trigger episodes is needed. Informing G+ individuals of the potential for deterioration in the setting of triggers, and the ongoing importance of general health is a helpful way of involving them in their care decisions. Further research of the G+P-group with high LVEF is needed to assess which patients may need less intensive surveillance, e.g., those with LVEF 50-55% or global longitudinal strain -16-18% should be more closely monitored than those with LVEF >60% on consecutive echocardiograms.

## Limitations

The main limitation of this study is the small sample size, and retrospective approach, which introduces the possibility of bias. Larger, multi-center, prospective studies are needed to explore these findings, however, conducting longitudinal studies in large cohorts with sufficient granularity to appraise these environmental factors is challenging. To avoid bias here, pre-specified criteria for environmental factors were used. In future studies, a blinded adjudication committee would be valuable to resolve complex cases. This cohort included small numbers of non TTN genotypes, some of which are expected to have more severe phenotypes. To account for this, a ‘severe disease gene’ cofactor was included in the analyses and did not change the observed outcomes. This was accounted for as a cofactor in the analyses. In the trajectory analysis of this study, a degree of reporter bias was unavoidable. Changes in LVEF were noted and correlated with changes in environmental factors, however, changes in environmental factors were not necessarily correlated with changes in LVEF. This was due to the complexity of accurately tracking changes in every environmental factor in each case. The use of reported LVEF in this study was potentially problematic as large variation can be seen between studies, vendors, and operators. An absolute LVEF change of 10% was considered a meaningful difference as this has previously been established as an acceptable cut-off for true change.^25^ In this study, LVEF was recorded from reports, and measurements were not standardized. Despite this, a comparison of LVEF values from clinical reports and a central laboratory showed overall good agreement. Additionally, patient follow-up in this study was variable and irregular and changes in LVEF may have occurred between follow-up time points that was not detected. A prospective, blinded study, with regular follow-up intervals, and using similar methodology is needed.

## Conclusions

Our data delineate a key role for environmental factors in the penetrance, trajectory, and outcomes of familial DCM. Personalized management strategies that incorporate risk factor surveillance could help to attenuate disease progression.

**Table 4.** Univariable and Multivariable Analyses of Adverse Events.

|  | Univariate |  |  | Multivariate |  |  |
| --- | --- | --- | --- | --- | --- | --- |
|  | OR | 95% CI | *P* | OR | 95% CI | *P* |
| Age at event (yr) | 1.00 | 0.97-1.04 | 0.79 | 1.01 | 0.97-1.07 | 0.56 |
| Sex (F) | 0.5 | 0.17-1.48 | 0.20 | 0.26 | 0.043-1.6 | 0.14 |
| Severe-phenotype gene | 3.18 | 0.21-3.18 | 0.76 | 1.53 | 0.20-11.42 | 0.67 |
| Baseline LVEF (%) | 1.18 | 1.12-1.25 | **<0.001** | 1.26 | 1.15-1.39 | **<0.001** |
| DCM-promoting factors (No.) | 1.74 | 0.83-3.64 | 0.14 | 1.91 | 0.35-10.51 | 0.45 |
| HF comorbidities (No.) | 1.88 | 1.15-3.01 | **0.014** | 4.9 | 1.73-14.08 | **0.004** |

Univariate and multivariate analysis using logistic regression with adverse event composite: cardiovascular death, heart transplant, heart failure hospitalisation or ventricular arrhythmias (ventricular fibrillation arrest, sustained ventricular tachycardia). Threshold for significance, *P* value <0.05.
CI = confidence interval; LVEF = left ventricular ejection fraction; OR = odd ratio; severe-phenotype gene = *BAG3*, *LMNA*, *RBM20*, *TNNT2*.

## Data Availability

Data can be made available by the authors upon reasonable request.

## Abbreviations

DCM: dilated cardiomyopathy
G+P+: genotype-positive, phenotype-positive
G+P-: genotype-positive, phenotype-negative
G-: genotype-negative
HF: heart failure
LP: likely-pathogenic
LV: left ventricular
LVEF: left ventricular ejection fraction
LVRR: left ventricular reverse remodelling
P: pathogenic
*TTN*tv: Titin truncating variant

